# Evidence for a serotonergic subtype of major depressive disorder: A NeuroPharm-1 study

**DOI:** 10.1101/2021.06.17.21258740

**Authors:** Kristin Köhler-Forsberg, Brice Ozenne, Elizabeth B. Landman, Søren V. Larsen, Asbjørn S. Poulsen, Vibeke H. Dam, Cheng-Teng Ip, Anders Jørgensen, Michal Meyer, Hans R. L. Eiberg, Claus Svarer, Martin B. Jørgensen, Vibe G. Frokjaer, Gitte M. Knudsen

## Abstract

Selective serotonin reuptake inhibitors (SSRIs) are the first line pharmacological treatment of Major Depressive Disorder (MDD), but only about half of patients benefit from it. Cerebral serotonin 4 receptor (5-HT_4_R) binding measured with positron emission tomography (PET) is inversely related to serotonin levels and can serve as a proxy for brain serotonin levels. We here determine if 5-HT_4_R differs between healthy and MDD individuals and if it is associated with successful outcomes of serotonergic treatment of MDD. We [^11^C]-SB207145 PET-scanned 100 (71 F) untreated patients with moderate to severe MDD and 91 (55 F) healthy controls; 40 patients were re-scanned after 8 weeks treatment. All patients started treatment with the SSRI escitalopram and were followed clinically after 1, 2, 4, 8 and 12 weeks. Treatment response was measured as change from baseline.

Before treatment, patients with MDD had 8% lower global 5-HT_4_R binding than controls (95%CI[-13.1%;-2.5%], p<0.001). Non-responders did not differ from controls, whereas remitters had 9% lower binding than controls ([-16.1%;-2.7%], p=0.004). Independent of treatment outcomes, patients reduced their neostriatal 5-HT_4_R binding (−9%, [-12.8%;-5.0%], p<0.001) after serotonergic intervention.

Overall, patients with MDD have lower cerebral 5-HT_4_R binding than controls, suggesting that 5-HT_4_R is a biomarker for MDD. The observation that SSRI treatment leads to reduced neostriatal 5-HT_4_R binding supports that the treatment does indeed increase brain 5-HT levels. Patients who remit to SSRIs have lower cerebral 5-HT_4_R prior to treatment than controls whereas non-responders do not differ. We propose that non-responders to SSRI’s constitute a subgroup with non-serotonergic depression.

ClinicalTrials.gov Identifier: NCT02869035

Registry name: Treatment Outcome in Major Depressive Disorder

URL: https://clinicaltrials.gov/ct2/show/NCT02869035?term=NCT02869035&draw=2&rank=1

## Introduction

Major depressive disorder (MDD) is one of the most frequent mental disorders worldwide^1^ and the demand for efficient and reliable treatments of MDD is high. The primary pharmacological treatment of MDD is selective serotonin reuptake inhibitors (SSRI) but about half of patients do not respond adequately to SSRI treatment.^2^ Currently, clinical practice is to make trial-and-error drug prescriptions which clearly has direct and adverse implications for those patients who are insufficiently treated. While the lack of predictability of antidepressant drug response remains a tremendous clinical challenge, it can be argued that one reason for inconsistent treatment response is due to patient heterogeneity within the broader diagnosis of MDD and that a better stratification is necessary to properly anticipate drug efficacy. For this purpose, access to biomarkers that allow for distinction between MDD subtypes would be highly valuable. Lack of response to drug treatment could potentially be due to different underlying pathophysiological mechanisms that happen to generate a clinical picture best described by a diagnosis of MDD. That is, identification of patient subgroups with different etiologies would enable a precision medicine approach where the treatment targets the specific neurobiological dysfunction.^3,4^ Additionally, identification of biomarkers that can identify patient subgroups who do not benefit from SSRIs could also be enormously helpful for optimizing clinical care and for future drug development programs.

Molecular neuroimaging with positron emission tomography (PET) and specific serotonergic radioligands is a highly sensitive method for direct investigation of the brain’s serotonergic transmitter system. The serotonin transporter (5-HTT) and the 5-HT_1A_ receptor (5-HT_1A_R) are the to date most studied serotonergic targets. PET studies of the 5-HTT have generated mixed outcomes of either none, decreased or even increased cerebral binding in MDD versus healthy controls.^5^ PET studies of the 5-HT_1A_R have also generated discrepant findings regarding the direction of 5-HT_1A_R modulation between MDD and healthy controls, depending on the quantification methodology used; the most pronounced difference was seen in the raphe nuclei and only in males.^6^ One study report that high raphe nuclei 5-HT_1A_R binding was associated with remission to escitalopram treatment 6-8 weeks later^7^ while others found higher 5-HT_1A_R orbital cortex binding in non-responders.^8^ Importantly, these PET studies typically include maximally 25 patients each, with some of the patients being medicated and or having significant comorbidity that could confound the interpretation.

The serotonin 4 receptor (5-HT_4_R) has been recognized as a new potential therapeutic target and both preclinical and human data support its involvement in MDD.^9^ The 5-HT_4_R is a G_s_ protein-coupled postsynaptic heteroreceptor, widely distributed in the brain.^10^ PET neuroimaging with the 5-HT_4_R radiotracer [^11^C]-SB207145 allows for studies of the receptor *in vivo*. In rodents, cerebral 5-HT_4_R binding is inversely related to changes in brain serotonin levels,^11–13^ and in a study of healthy individuals PET-scanned before and after three weeks of SSRI or placebo found reduced 5-HT_4_R binding, also in support of an inverse relation between 5-HT_4_R and cerebral serotonin levels.^14^ Some evidence for cerebral 5-HT_4_R being a trait biomarker for MDD comes from studies of healthy people with first-degree relatives with the disorder: having a family history of MDD is associated with lower striatal 5-HT_4_R binding, and the more relatives with MDD, the lower the striatal and limbic 5-HT_4_R binding.^14^

Here, we applied a naturalistic study design and enrolled 100 pharmacologically untreated patients with moderate to severe MDD; they were assessed clinically and investigated at baseline with [^11^C]-SB207145 PET and magnetic resonance imaging (MRI) neuroimaging before they were started on standard SSRI treatment. To take into account that the clinical effect of SSRIs can be delayed for weeks,^15^ we regularly assessed the patients clinically for up to 12 weeks. After 8 weeks of treatment, 43 of the patients were rescanned with PET and MRI. The aims of our study were to investigate if:

a. patients with MDD differ in cerebral 5-HT_4_R binding at baseline compared to healthy controls
b. cerebral 5-HT_4_R binding in patients with MDD is associated with remission within 8 weeks after starting SSRI treatment
c. remitted patients with MDD show larger reduction in their cerebral 5-HT_4_R binding than non-responders.

## Subjects and methods

One hundred antidepressant-free outpatients with moderate to severe MDD were recruited from the mental health system in the capital region of Denmark and included in a non-randomized, 12-week longitudinal, open clinical trial where they received standard antidepressant drug treatment. All participants provided written informed consent prior to inclusion and recruitment was performed by a trained study physician. The study protocol was approved by all relevant authorities (the Health Research Ethics Committee of the Capital Region of Denmark (H-15017713), the Danish Data Protection Agency (04711/RH-2016-163) and Danish Medicines Agency (EudraCT-2016-001626-34)) and registered as a clinical trial before initiation (NCT02869035). The study was monitored by an external good clinical practice unit from the capital region of Copenhagen, Denmark. Patients between 18–65 years of age and with a Hamilton Depression Rating Scale 17 items (HAMD_17_)^16^ score >17 were included. Patients were screened with the Mini International Neuropsychiatric Interview^17^ and the diagnosis was confirmed by a specialist in psychiatry. Exclusion criteria were: use of antidepressant medicine within the last two months; duration of the present depressive episode exceeding two years; more than one attempt with an antidepressant treatment in the current episode; previous non-response or known contraindications to an SSRI drug, other primary axis I psychiatric disorder; alcohol/substance abuse or dependence; severe somatic illness; insufficient language skills in Danish; acute suicidal ideation or psychosis; current or planned pregnancy or breast feeding; use of medical treatment affecting CNS (e.g., metoclopramide, ondansetron, serotonergic drugs for migraine, clonidine); contraindications to PET/MRI scans; history of severe brain injury or significant cognitive impediments. Ninety-one healthy controls were included for baseline comparisons and recruited either from our quality-controlled repository^18^ or from an online recruitment site, meeting the same in-and exclusion criteria as the patients (except no past or present psychiatric disorders). The healthy controls matched the patients’ age and sex as closely as possible. The method and study design are described in detail elsewhere.^19^

### Study assessments for participants and treatment course for patients

Before inclusion, medical history and prior medical treatment was assessed. All participants underwent somatic and psychiatric screening, urine screening for pregnancy or toxicology, and routine blood tests. At baseline, participants were brain scanned with MRI and [^11^C]-SB207145 PET and 43 of the patients were PET and MRI rescanned at week 8. After completion of the baseline program, patients started antidepressant treatment with escitalopram, individually adjusted to 10-20 mg daily depending on response and side effects. Clinical treatment response was monitored after 1, 2, 4, 8 and 12 weeks of treatment by face-to face visits and HAMD_17_ and HAMD_6_ ratings^20^. Regular co-ratings between study investigators were implemented. Patients with intolerable side effects or < 25 % reduction from baseline in HAMD_6_ at week 4 were offered to switch to the serotonin-norepinephrine reuptake inhibitor, duloxetine, individually adjusted (30-120 mg daily). Serum concentration of escitalopram or duloxetine was determined at week 8.

### Clinical outcome measures

The primary clinical outcome measure was change in HAMD_6_ from baseline to week 8. Remitters were defined as having a ≥ 50 % reduction in HAMD_6_ at week 4 (early responders) and HAMD_6_ score < 5 at week 8. Non-responders had < 25% reduction in HAMD_6_ at week 4 (early non-responder) and < 50% reduction in HAMD_6_ at week 8. Patients in between these categories were categorized as intermediate responders. As a secondary clinical outcome measure, we used relative percentage change in HAMD_6_ (rΔHAMD_6_) from baseline to week 2, 4, 8 and 12.

### PET and MRI procedure

PET/MRI acquisition, pre-processing and PET quantification was performed as previously described.^19^ Briefly, PET images were acquired during a 120 minutes dynamic scan using a high-resolution research tomography Siemens PET scanner (CTI/Siemens, Knoxville, TN, USA) after intravenous injection of [^11^C]-SB207145. All patients and 53 controls were scanned with a Siemens 3-Tesla Prisma and 38 controls with a Siemens Magnetom Trio 3-Tesla MRI scanner. 3D T1-weighted MRI was co-registered to PET images to obtain structural information. PET scans were motion corrected using the Air 5.2.5 method.^21^ PVE-lab was used to extract region of interest (ROIs),^22^ delineated on the individuals’ MRI. The mean tissue time activity for hemisphere-averaged grey matter volumes was used for kinetic modeling with cerebellum (excluding vermis) as a reference region.^23^ The calculated non-displaceable binding potential (BP_ND_) served as an outcome measure for the 5-HT_4_R binding.

### Statistics

We included 100 patients to reach a statistical power of 0.8 for detection of a 7% difference in BP_ND_ between remitters and non-responders, with an expected drop-out rate of 20%. For the descriptive statistics, the p-value was computed using Fisher’s exact t-test for categorical and Mann Whitney U-test for continuous variables respectively. For the primary analysis, we used a latent variable model (LVM) to test for global and regional differences in (i) baseline 5-HT_4_R BP_ND_ between patients and controls, (ii) baseline 5-HT_4_R BP_ND_ between remitters, non-responder, and controls, and (iii) change in BP_ND_ between baseline and follow up (ΔBP_ND_) and whether ΔBP_ND_ differed between remitters and non-responders. We included neocortex, hippocampus, caudate nucleus and putamen^22^ as regions of interest in the LVM because of their relevance in mood disorders.

Secondary analyses included testing with LVM (ii’) for an association between baseline BP_ND_ and rΔHAMD_6_ (ii’’), for a difference in baseline BP_ND_ between early responder, early non-responder, and healthy controls, and (iii’) for an association between ΔBP_ND_ and rΔHAMD_6_. In order to assess the data by more commonly used statistics, analyses (i), (ii), (ii’), and (ii’’) were also performed using multiple linear regressions one for each brain region (neocortex, a limbic region and neostriatum).

Beyond testing for associations, we also evaluated the prognostic value of 5-HT_4_R binding in baseline for the outcome of antidepressant treatment. For each brain region, the prognostic value of a low 5-HT_4_R BP_ND_ was assessed using the area under the ROC curve (AUC). Here the AUC is the probability that a remitter has a lower 5-HT_4_R BP_ND_ than a non-responder, 0.5 indicating no prognostic value. The positive predictive value (PPV) and negative predictive value (NPV) of the 5-HT_4_R BP_ND_ were assessed by dichotomizing 5-HT_4_R BP_ND_ at the threshold which maximized the Youden-Index. All analyses were adjusted for age, sex, injected SB207145 (mass/kg), the 5-HT transporter polymorphism (5-HTTLPR) genotype (L_A_L_A_ or non-L_A_L_A_), and MR-scanner,^24–27^ except within subject rescan-analyses (iii) and (iii’) that were only adjusted for the difference in injected SB207145 (mass/kg) between baseline and week 8. When using LVMs, the covariates were included in the measurement model. 5-HT_4_R BP_ND_ values were log-transformed. When using LVMs, score tests were used to detect model misspecifications and additional parameters were included until no misspecification could be detected. Missing data in analysis (ii) were handled using complete case analysis. We also adapted an alternative approach where missing values in the primary clinical outcome were imputed based on the clinical outcome at week 4. Nine patients left the study prematurely: those leaving due to early remission were classified as remitters; those leaving because of side effects or suicidality as non-responders. Inverse probability weighting was used to handle other types of dropout using baseline covariates as predictors of dropout. Secondary analyses were performed using complete case analysis. Reported p-values and 95% confidence intervals were two-sided. When performing tests across several brain regions we adjusted p-values (p.adj) and confidence intervals using a single-step Dunnett procedure.^28^ All analyses were performed in R.

## Results

Patients were recruited and followed between the Aug 15, 2016 to April 17, 2019. Supplementary Figure 1 shows the CONSORT diagram. Demographics, clinical profile and tracer data (Table 1) showed that patients and controls were comparable, except for injected mass/kg and a minor difference in education. We included 91 patients for baseline analyses and 78 in the longitudinal analyses; of the latter, 22 were remitters and 13 non-responders after 8 weeks, and 34 were early responders and 14 early non-responders after 4 weeks. Supplementary Table 1 describes baseline psychopathological profile for non-responders and remitters. Six patients switched to duloxetine before week 8. Re-scan data was obtained from 12 remitters, five non-responders and 23 responders. No serious adverse events occurred during the study. Remission rate was 48% at week 12 according to remission-criteria used in, e.g., STAR*D study (HAMD_17_ ≤ 7)^29^ and comparable to similar clinical trials.^30^

**Table 1.** Clinical profile, demographic and radiotracer data for patients with MDD and controls at baseline. BMI: body mass index. HAMD_17/6_: Hamilton depression rating scale 17 or 6 items. MDI: Major depressive inventory. NA: not applicable. ^a^ p-value computed using a Fisher’s exact t-test, ^b^ p-value computed using a Mann Whitney U-test.

|  | Patients with MDD |  |  | Healthy controls |  |  | p-value <sup>a</sup> |
| --- | --- | --- | --- | --- | --- | --- | --- |
|  |  | n | % |  | n | % |  |
| Sex | Female | 65 | 71.4 |  | 55 | 60.4 | 0.16 |
|  | Male | 26 | 28.6 |  | 36 | 39.6 |  |
| 5-HTTLPR genotype | L<sub>A</sub>L<sub>A</sub> | 26 | 28.6 |  | 27 | 29.7 | 1 |
|  | Non-L<sub>A</sub>L<sub>A</sub> | 65 | 71.4 |  | 64 | 70.3 |  |
|  | Range | n | Mean (SD) | Range | n | Mean (SD) | p-value <sup>b</sup> |
| Age (years) | 18.3-57.3 | 91 | 27.1 (8.2) | 19.2-60.1 | 91 | 27.1 ± 8.0 | 0.57 |
| Years of education | 5-12 | 76 | 11.6 (1.1) | 9-12 | 91 | 11.9 (0.5) | **0.003** |
| BMI (kg/m<sup>2</sup>) | 17.1-45.1 | 91 | 24.5 (5.6) | 18.3-36.9 | 91 | 23.6 (3.1) | 0.96 |
| HAMD<sub>17</sub> | 18-31 | 91 | 22.9 (3.4) | NA |  | NA | NA |
| HAMD<sub>6</sub> | 7-17 | 91 | 12.3 (1.6) | NA |  | NA | NA |
| MDI | 16-50 | 89 | 34.7 (7.2) | 0-18 | 91 | 5.6 (4.2) | **< 0.001** |
| Injected dose (MBq) | 263.0-615.0 | 91 | 577.4 (56.0) | 226-617 | 91 | 569.4 (76.3) | 0.20 |
| Injected mass/kg (µg/kg) | 0.004-0.082 | 91 | 0.013 (0.015) | 0.003-0.07 | 91 | 0.017 (0.015) | **0.028** |
| Cerebellum, area under curve (kBq/ml) | 3.9-17.8 | 91 | 10.3 (2.6) | 3.2-16.2 | 85 | 10.3 (2.5) | 0.75 |

We found 7-8% lower regional BP_ND_ in untreated patients with MDD compared to controls (p <0.001), (Figure 1 and Supplementary Figure 2). Linear regression models generated the same outcome (Supplementary Table 2). Since BP_ND_ in the caudate nucleus and putamen were especially correlated, these regions were pooled into “neostriatum” for the subsequent analyses.

**Figure 1.**
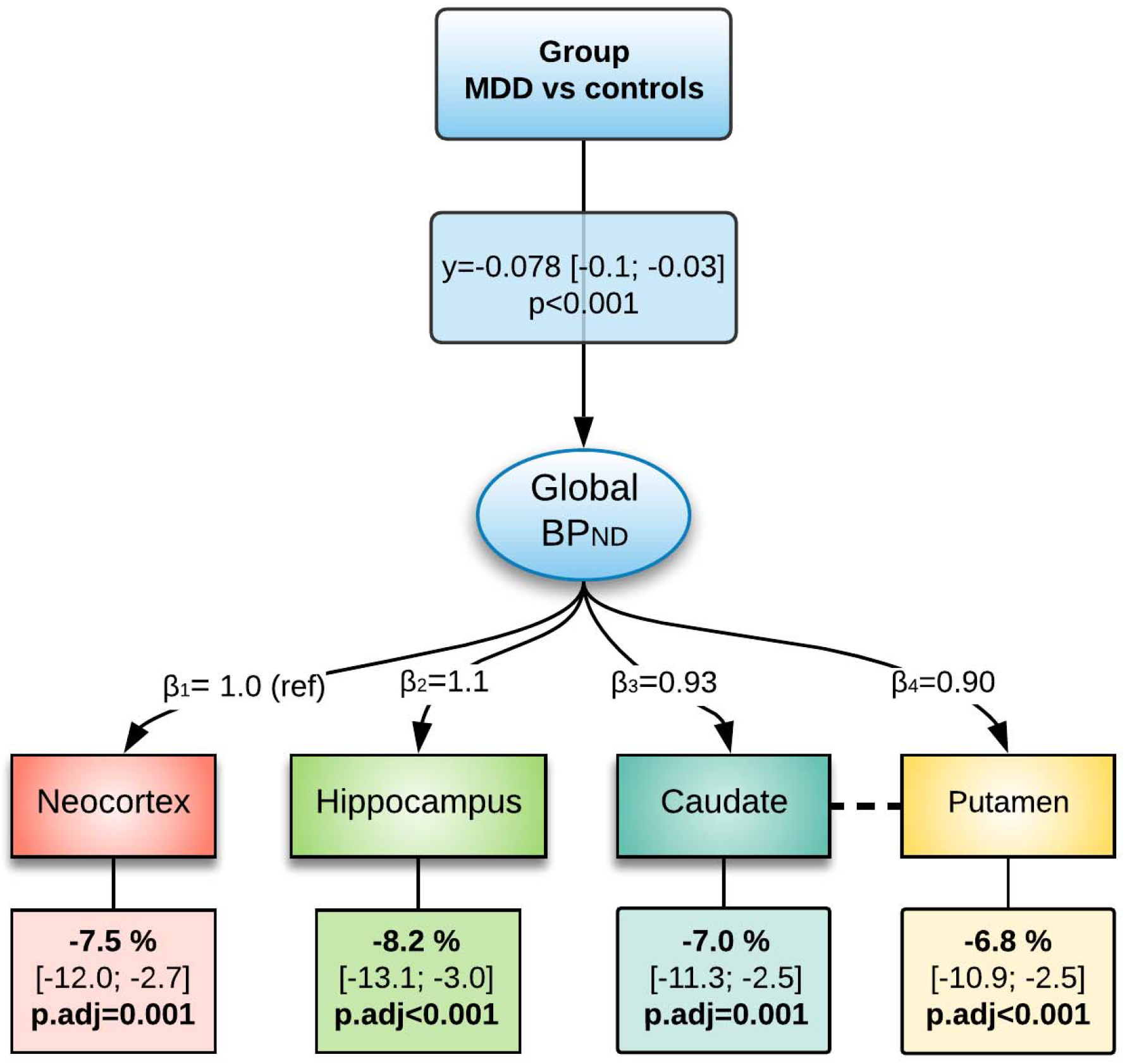
Estimated latent variable model for the 5-HT4R binding in untreated patients with MDD and controls. γ is the effect of group-status on the global (log-transformed) BP_ND_, β is the loading, the dashed line indicates additional shared correlations between caudate nucleus and putamen. The lower boxes indicate, for each brain region, the percentage difference in baseline 5-HT_4_R binding between MDD and controls (p-values and confidence intervals are adjusted for 4 comparisons). Age, sex, 5-HTTLPR gene-status, MR-scanner type and injected mass/kg are included as covariates in the model.

Global BP_ND_ was lower in remitters than in controls (p=0.004, Table 2), with 8-10% lower binding in neocortex (Figure 2), hippocampus and neostriatum. Supplementary Figure 3 displays the baseline BP_ND_ for patients according to clinical response group and controls. There was no statistically significant difference in global BP_ND_ between non-responders and controls (p=0.31) or between remitters and non-responders (p=0.18). Handling missing data using a combination of imputation and inverse probability weighting lead to estimates and conclusions that were similar to the complete case analysis (Supplementary Table 3). Response categories at week 4 (Table 2) showed 8-10% lower BP_ND_ in early responders than in controls (p=0.002), and 7-9% lower BP_ND_ in early responders compared to early non-responders (p=0.046). There was no difference between early non-responders and controls (p=0.79). Similar results were found when using multiple linear regression (Supplementary Table 4). Further, we found a correlation between baseline BP_ND_ and rΔHAMD_6_ at week 4 (p=0.03), but not at week 8 (p=0.98) using LVM. Univariate analysis identified the correlation in neocortex at week 4 only (Supplementary Table 5).

**Table 2.** Cerebral 5-HT_4_R binding in controls and in MDD, according to treatment response at week 4 and 8. The p-values refer to the testing of BP_ND_ between two groups across all regions. The last three rows display the region-specific difference in BP_ND_ between two groups with confidence interval, corrected for three comparisons (i.e. across the three regions). All estimates originate from the latent variable model.

|  | Early responder vs. controls | Early non-responder vs. controls | Early responder vs. early non-responder | Remitter vs. Control | Non-responder vs. Control | Remitter vs. Non-responder |
| --- | --- | --- | --- | --- | --- | --- |
| **n** | 34 vs. 91 | 14 vs. 91 | 34 vs. 14 | 22 vs. 91 | 13 vs. 91 | 22 vs. 13 |
| **Week** | 4 | 4 | 4 | 8 | 8 | 8 |
| **Global effect** |  |  |  |  |  |  |
| **p** | **0.002** | 0.79 | **0.046** | **0.004** | 0.31 | 0.18 |
| **Regional effect** |  |  |  |  |  |  |
| **Neocortex** | -8.96%<br>[-14.63; -2.91] | -1.03%<br>[-8.37; 6.9] | -8.01%<br>[-15.61; 0.27] | -9.5%<br>[-15.85; -2.66] | -3.89%<br>[-11.16; 3.98] | -5.84%<br>[-14.04; 3.16] |
| **Hippocampus** | -10%<br>[-16.27; -3.25] | -1.16%<br>[-9.34; 7.77] | -8.94%<br>[-17.34; 0.31] | -9.92%<br>[-16.54; -2.77] | -4.07%<br>[-11.65; 4.17] | -6.1%<br>[-14.65; 3.31] |
| **Neostriatum** | -7.68%<br>[-12.6; -2.47] | -0.88%<br>[-7.17; 5.84] | -6.86%<br>[-13.45; 0.23] | -8.19%<br>[-13.76; -2.27] | -3.34%<br>[-9.64; 3.4] | -5.02%<br>[-12.16; 2.7] |

**Figure 2.**
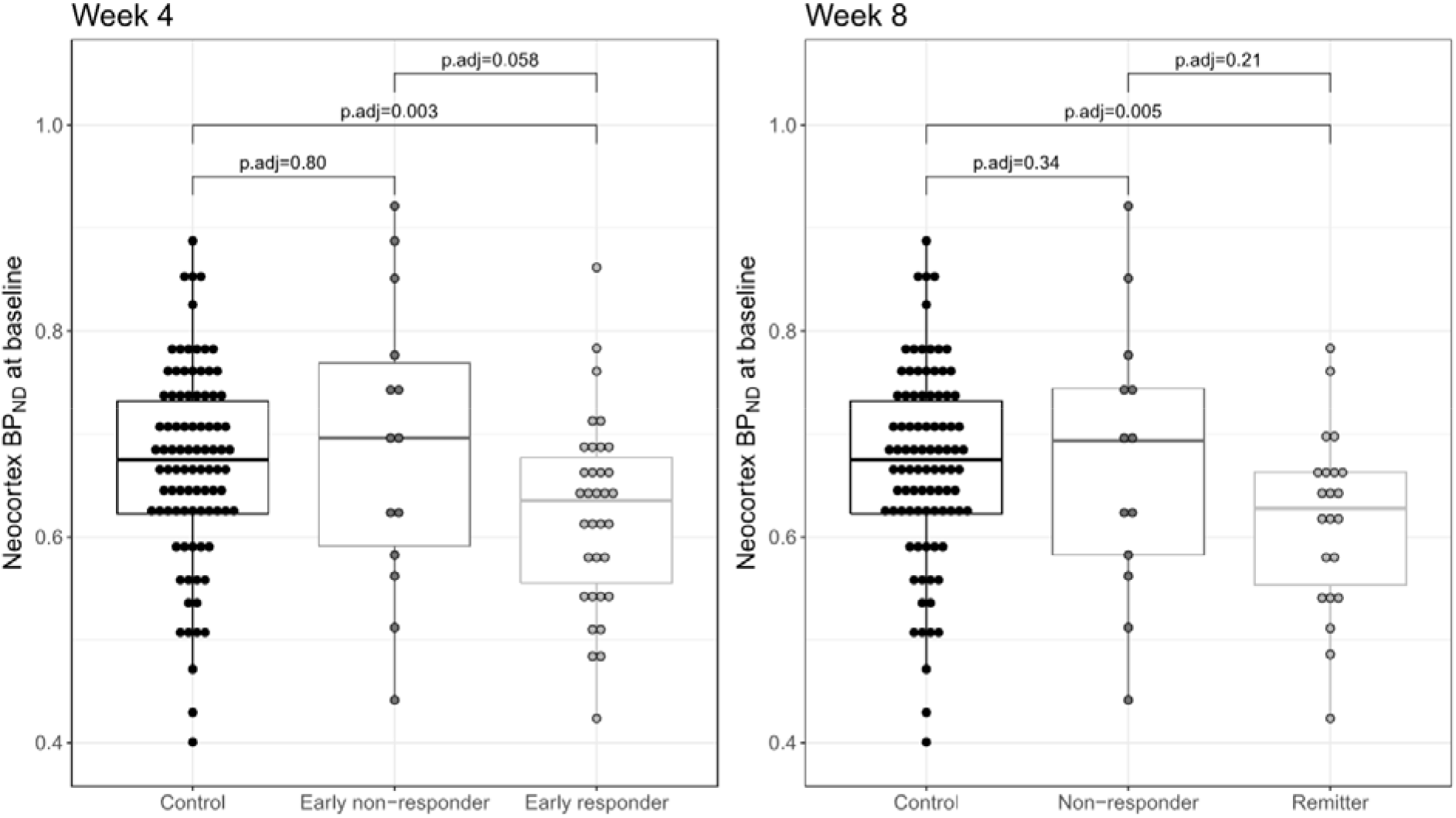
Scatter plot of 5-HT_4_R baseline binding in neocortex for healthy controls and patients with MDD according to clinical outcome at week 4 and 8. Week 4: Controls (n=91), early responders (n=34), and early non-responders (n=14). Week 8: Controls (n=91), remitters (n=22), and non-responders (n=13). P-values originate from the latent variable model and were adjusted for 3 comparisons.

Regional baseline BP_ND_ did not show prognostic power for identifying non-responders from remitters: neocortex (AUC: 0.63 [0.43; 0.84], p=0.20), limbic region (AUC: 0.57 [0.35; 0.79], p=0.54), neostriatum (AUC: 0.57[0.36; 0.77], p=0.52). Based on the Youden index, the regional baseline BP_ND_ was dichotomized at 0.69 (neocortex), 3.76 (neostriatum), and 0.90 (limbic). The estimated PPV were, respectively, 0.76 [0.55; 0.91], 0.72 [0.51; 0.88], and 0.72 [0.53; 0.87] and the estimated NPV were, respectively 0.70 [0.35; 0.93], 0.60 [0.26; 0.88], and 0.83 [0.36; 1]. This can be compared to a classifier using only the observed prevalence of remitters: classifying 63% of the patients as remitters and the rest as non-responders would lead to a PPV of 0.63 and a NPV of 0.37. The predictive values for various response status using baseline 5-HT_4_R binding are shown in Supplementary Table 6.

Eight weeks after initiating SSRI treatment, patients showed a decrease in global BP_ND_ compared to baseline (p<0.001, LVM-model, N=40). At a regional level, the decrease in BP_ND_ constituted 9.0% [-12.8%; −5.0%] in neostriatum (p.adj<0.0001) but no significant change was seen in neocortex (−1.4% [-6.2%; 3.6%], p.adj=0.79) or hippocampus (−1.7% [-7.5%; 4.5%], p.adj=0.80) (Figure 3 and Supplementary Figure 4). The decline was not associated with categorical response at week 8 (p=0.60) or rΔHAMD_6_ (p=0.74).

**Figure 3.**
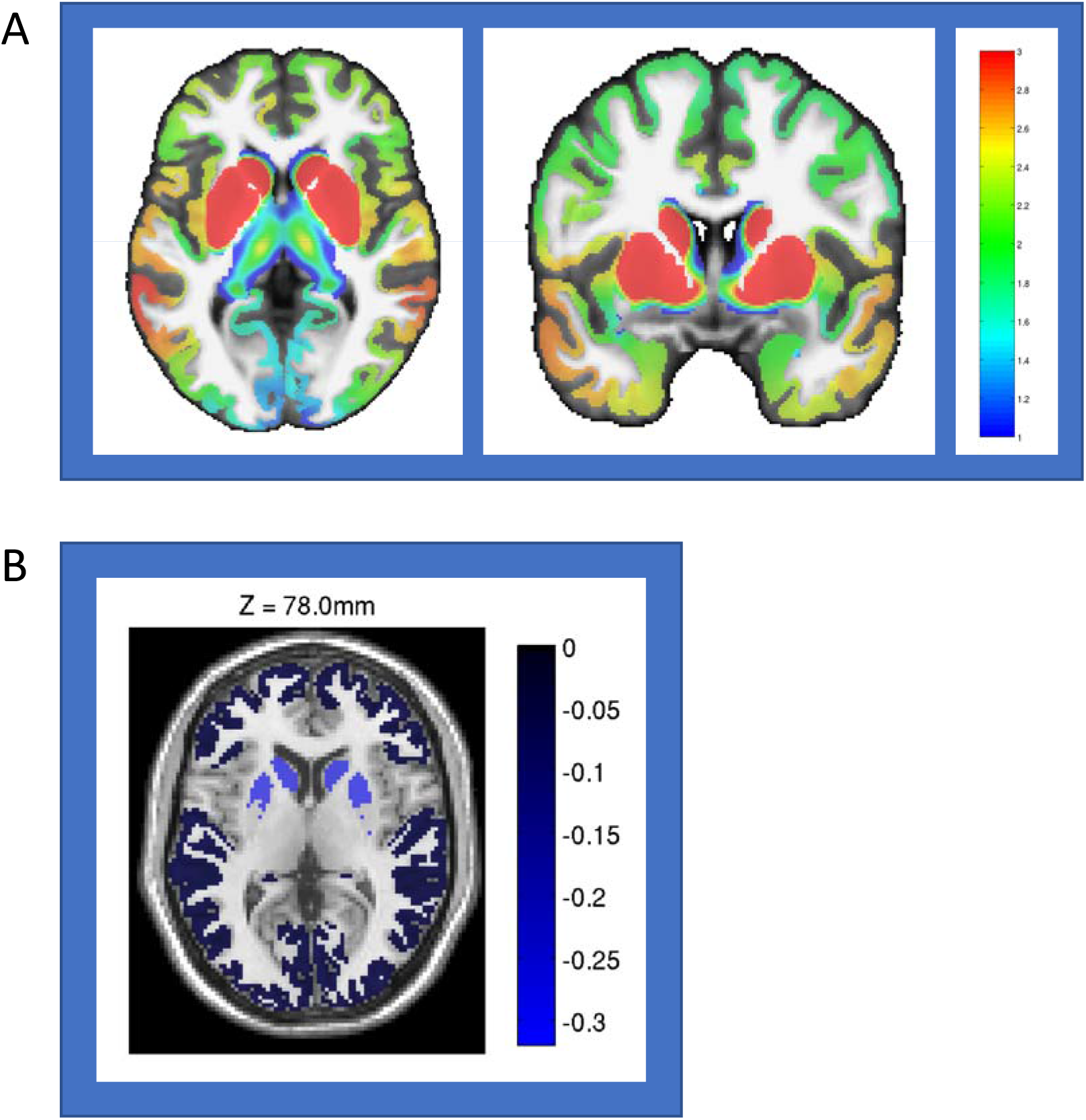
**Panel A**. Average density maps (pmol/ml) for the 5-HT_4_R at baseline in patients with MDD (n=91). Atlas used from Beliveau and colleagues (2017).^10^ **Panel B**. Difference in mean 5-HT_4_R binding from baseline and rescan in patients (N=40). Regions of interest for the latent variable model analyses (neocortex, hippocampus and neostriatum) are shown. The post-SSRI effect was most prominent in neostriatum (lighter blue).

## Discussion

In this to date largest single clinical PET trial investigating the serotonin system in MDD, we show that antidepressant-free patients with a moderate to severe major depressive episode on average have 7-8% lower cerebral 5-HT_4_R binding than healthy controls. Intriguingly, patients who remit after 4 or 8 weeks of serotonergic medication have 8-10% lower cerebral 5-HT_4_R baseline binding whereas non-responders do not differ from controls. When patients were PET-rescanned 8 weeks after starting SSRI treatment, their striatal 5-HT_4_R binding had decreased, irrespective of the clinical treatment outcome. These results support the notion that only a subgroup of patients with MDD have a serotonergic dysfunction and that accordingly patients within this subgroup are effectively treated with SSRI.

Our finding of abnormally low 5-HT_4_R binding in the subgroup of unmedicated MDD patients that remit on SSRI treatment could constitute a trait or a state feature. A previous study reported that the more first degree relatives with MDD a healthy individual has, the lower is striatal 5-HT_4_R binding,^31^ and it was suggested that low 5-HT_4_R binding could be a trait marker for increased risk of MDD, possibly reflecting increased cerebral serotonin levels that ensured euthymia. Since we here find a lower global 5-HT_4_R binding specifically in patients that remit in response to SSRI treatment, it seems less likely that low 5-HT_4_R binding is a *general* trait marker for unmedicated MDD. With the observed inverse relation between 5-HT_4_R binding and cerebral serotonin levels, ^11–14^ one interpretation is that already prior to treatment, remitters have higher brain serotonin levels. Increased serotonin levels could be the brain’s attempt to maintain euthymia, and addition of serotonergic acting drugs increases serotonin levels sufficiently for remission to occur. Alternatively, or in combination, patients responding to SSRIs may be genetically predisposed for low cerebral 5-HT_4_R density.

Our observation that striatal 5-HT_4_R binding decreases in response to increased serotonin levels also in patients with MDD (Supplementary Figure 5) is consistent with observations in preclinical studies and in healthy indivdiuals^11–14^. After 8 weeks of treatment, we found across response-groups a 9% decrease in neostriatum 5-HT_4_R binding, suggesting that it was not a failure of the drug to affect the brain serotonin levels that explained a poor clinical drug response. Interestingly, whereas the reduction in 5-HT_4_R binding seen after serotonergic treatment was specific for neostriatum, differences between patients with MDD and controls showed a global effect across all brain regions. The regional difference in the rescan data could be due to the drug intervention having a specific effect by increasing serotonin in neostriatum^13^ which together with thalamus is massively innervated by serotonergic fibers and has among the highest density of serotonin transporters.^10^

Short-term administration of 5-HT_4_R agonists to rodents generates rapid antidepressant/anxiolytic-like behavior,^32,33^ hippocampal neurogenesis,^34^ prophylactic antidepressant and anxiolytic characteristics^35^ and the first translational study recently confirmed that administration of the 5-HT_4_R agonist prucalopride enhances memory effects in healthy volunteers.^36^ It remains to be tested in clinical trials if 5-HT_4_R agonists could constitute a new therapeutic target for the MDD serotonergic subtype patients. Our data also opens for an interesting possibility of identification of a distinct biological subtype within MDD with a “non-serotonergic”-related depression; such a subgroup would be amenable for investigation of non-serotonergic drug effects.

In conclusion, we here provide novel support that MDD patients with a primary serotonergic dysfunction constitute a subgroup where SSRI/SNRI treatment is particularly effective. Neuroimaging of the 5-HT_4_R can thus be regarded as a biomarker that aids to identify subgroups of patients with MDD (e.g., non-serotonergic related depression) which can guide future clinical trials in MDD and enable future precision medicine approaches.

## Data Availability

Data can be shared upon request and in compliance with GDPR regulations. www.nru.dk

## Acknowledgements

We thank all participants and their relatives for taking part in this study. We gratefully acknowledge all investigators involved, collaborating general practitioners, the Center for Referral and Diagnostics, Mental Health Services, Capital Region of Copenhagen and other collaborators who helped throughout the study. We especially thank Lone Ibsgaard Freyr, Bente Dall, Gerda Thomsen, Svitlana Olsen, Agnete Dyssegaard, Arafat Nasser, Ida Marie Brandt and Anna Maria Florescu for their excellent laboratory and technical assistance. Economic support was granted from the Innovation Fund Denmark, The Lundbeck Foundation alliance BrainDrugs (R279-2018-1145), Research Fund of the Mental Health Services - Capital Region of Denmark, Independent Research Fund Denmark, G.J. Foundation, Research Council of Rigshospitalet, Augustinus Foundation, Savværksejer Jeppe Juhl og hustru Ovita Juhls Mindelegat, and the H. Lundbeck A/S. H. The funders of the study had no role in study design, data collection, data analysis, data interpretation, or writing of the report. The corresponding author had full access to all the data in the study and had final responsibility for the decision to submit for publication.

