## Supplementary material for "Evidence for a serotonergic subtype of major depressive disorder: A NeuroPharm-1 study"

#### Tables 1-6

| | Remitter (n=22) | | Non-responder (n=13) | |
|---|---|---|---|---|
| | *Range* | *Mean (SD)* | *Range* | *Mean (SD)* |
| **$HAMD_{17}$ baseline** | 18-29 | 22·9 (3·0) | 18-27 | 21·5 (2·1) |
| **$HAMD_6$ baseline** | 8-14 | 11·9 (1·5) | 7-14 | 11·2 (1·8) |
| **$HAMD_6$ items** | | | | |
| - **Depressed mood** | 2-4 | 2·7 (0·6) | 2-4 | 3·1 (0·5) |
| - **Feelings of guilt** | 0-3 | 1·6 (0·7) | 0-3 | 1·5 (0·8) |
| - **Work and activities** | 1-4 | 2·3 (0·7) | 2-3 | 2·3 (0·5) |
| - **Retardation** | 0-3 | 1·2 (0·8) | 0-2 | 0·5 (0·8) |
| - **Anxiety (psychic)** | 1-4 | 2·5 (0·7) | 0-3 | 2 (0·8) |
| - **Somatic symptoms (general)** | 0-2 | 1·6 (0·7) | 0-2 | 1·8 (0·6) |
| **MDI baseline** | 16-50 | 32·9 (9·0) | 29-45 | 35·9 (5·9)* |

**Supplementary Table 1. Descriptive table of baseline psychopathology in non-responders and remitters.** $HAMD_6$ items are specified. *One patient failed to fill out MDI at baseline. MDI: Major Depression Inventory. $HAMD_{17/6}$: Hamilton Depression Rating Scale 17 and 6 items. Group differences tested using a Mann Whitney U-test.

| ROI | % change in $BP_{ND}$ (MDD vs control) | Standard error | 95% CI | p.adj |
|---|---|---|---|---|
| **Neocortex** | -8·86 | 2·31 | -14·00; -3·41 | <0·001 |
| **Limbic region** | -6·53 | 2·14 | -11·42; -1·37 | 0·011 |
| **Neostriatum** | -6·21 | 2·16 | -11·15: -0·99 | 0·017 |

**Supplementary Table 2. Regional percentage difference in 5-$HT_4$R binding in MDD versus controls at baseline.** Estimated by multiple linear regressions adjusted for age, sex, 5-HTTLPR status, injected tracer (mass/kg) and MR scanner type. P-values are adjusted for multiple comparison.

| | **Remitter vs. Control (ref)** | **Non-responder vs. Control (ref)** | **Remitter vs. non-responder (ref)** |
|---|---|---|---|
| n | 23 vs. 91 | 16 vs. 91 | 23 vs. 16 |
| week | 8 | 8 | 8 |
| p | **0·01** | 0·34 | 0·22 |
| Neocortex | -8·11% [-14·33; -1·42] | -3·31% [-9·96;3·83] | -4·96% [-12·62;3·37] |
| Hippocampus | -8·39% [-14·82; -1·47] | -3·42% [-10·30;3·98] | -5·14% [-13·05;3·50] |
| Neostriatum | -6·95% [-12·37; -1·20] | -2·83% [-8·55;3·26] | -4·25% [-10·87;2·87] |

**Supplementary Table 3. Cerebral 5-HT$_4$R binding in controls and in MDD according to treatment outcome in week 8 after accounting for missing data.** The p-values reflect the difference in $BP_{ND}$ between two groups across all regions. The last three rows display the region-specific difference in binding between two groups with confidence intervals, corrected for 3 comparisons (i.e. across the 3 regions). All estimates originate from the latent variable model after imputation and inverse probability weighting.

| | Early responder vs. control (ref) | Early non-responder vs. control (ref) | Early responder vs. early non-responder (ref) | Remitter vs. control (ref) | Non-responder vs. control (ref) | Remitter vs. non-responder (ref) |
|---|---|---|---|---|---|---|
| **n** | 34 vs 91 | 14 vs 91 | 34 vs 14 | 22 vs 91 | 13 vs 91 | 22 vs 13 |
| **Week** | 4 | 4 | 4 | 8 | 8 | 8 |
| **Neocortex** | -7·95% -14·49; -0·91] | 2·16% [-8·07;13·53] | -9·9% [-20·21;1·73] | -8·52% [-16·37;0·08] | -0·06% [-10·55; 11·65] | -8·46% [-20·13;4·92] |
| **p.adj** | **0·025** | 0·85 | 0·10 | 0·052 | 1·00 | 0·25 |
| **Limbic region** | -4·48% [-11·27;2·84] | 2·79% [-7·63;14·38] | -7·07% [-17·92;5·21] | -4·08% [-11·81;4·32] | 0·13% [-9·74;11·09] | -4·21% [-15·72;8·87] |
| **p.adj** | 0·28 | 0·77 | 0·30 | 0·44 | 1·00 | 0·67 |
| **Neostriatum** | -4·92% [-11·14;1·73] | 0·93% [-8·47;11·3] | -5·8% [-15·91;5·52] | -5·25% [-13·01;3·2] | -1·42% [-11·35;9·61] | -3·89% [-15·76;9·67] |
| **p.adj** | 0·17 | 0·97 | 0·37 | 0·27 | 0·94 | 0·73 |

**Supplementary Table 4.** Association between baseline 5-HT$_4$R binding and categorical response group week 4 and week 8 using linear regression. Adjusted for age, sex, 5-HTTLPR status, MR-scanner type and injected tracer (mass/kg).

| Region | Week | Partial correlation | Confidence Interval | p | p.adj |
|---|---|---|---|---|---|
| Neocortex | 2 | 0·19 | [-0·04;0·41] | 0·10 | 0·18 |
| | 4 | 0·31 | [0·09;0·52] | **0·005** | **0·01** |
| | 8 | 0·09 | [-0·14;0·31] | 0·44 | 0·64 |
| | 12 | 0·05 | [-0·19;0·28] | 0·70 | 0·91 |
| Limbic region | 2 | 0·08 | [-0·14;0·30] | 0·47 | 0·71 |
| | 4 | 0·22 | [0·003;0·44] | **0·047** | 0·09 |
| | 8 | 0·01 | [-0·21;0·24] | 0·90 | 1·00 |
| | 12 | -0·02 | [-0·24;0·21] | 0·88 | 0·99 |
| Neostriatum | 2 | 0·06 | [-0·17;0·28] | 0·62 | 0·86 |
| | 4 | 0·14 | [-0·08;0·36] | 0·21 | 0·35 |
| | 8 | -0·03 | [-0·26;0·20] | 0·79 | 0·96 |
| | 12 | 0·03 | [-0·20;0·25] | 0·83 | 0·98 |

**Supplementary Table 5.** Correlation between baseline 5-$HT_4R$ binding and percentage change in $HAMD_6$ at week 2-12, using partial correlation correction. Covariates: age, sex, 5-HTTLPR status and injected tracer (mass/kg).

| Outcome | Region | AUC | 95% CI | p-value |
|---|---|---|---|---|
| Remission vs. intermediate response and non-response | Neocortex | 0·49 | [0·36;0·63] | 0·93 |
| Non-response vs. intermediate response and remission | Neocortex | 0·65 | [0·46;0·84] | 0·11 |
| $HAMD_6 \leq 5$ (week 8) | Neocortex | 0·51 | [0·38;0·64] | 0·87 |
| Relative change in $HAMD_6 \leq 5$ (week 8) | Neocortex | 0·51 | [0·38;0·65] | 0·85 |
| Remission vs. intermediate response and non-response | Neostriatum | 0·48 | [0·34;0·62] | 0·79 |
| Non-response vs. intermediate response and remission | Neostriatum | 0·59 | [0·42;0·76] | 0·30 |
| $HAMD_6 \leq 5$ (week 8) | Neostriatum | 0·48 | [0·35;0·61] | 0·74 |
| Relative change in $HAMD_6 \leq 5$ (week 8) | Neostriatum | 0·54 | [0·41;0·67] | 0·58 |
| Remission vs. intermediate response and non-response | Limbic region | 0·46 | [0·33;0·59] | 0·53 |
| Non-response vs. intermediate response and remission | Limbic region | 0·59 | [0·39;0·80] | 0·37 |
| $HAMD_6 \leq 5$ (week 8) | Limbic region | 0·49 | [0·36;0·62] | 0·85 |
| Relative change in $HAMD_6 \leq 5$ (week 8) | Limbic region | 0·49 | [0·35;0·63] | 0·90 |

**Supplementary Table 6**. Predictive value for various response status using baseline 5-$HT_4R$ binding in neocortex, neostriatum and the limbic region. P-value is not adjusted.

### Supplementary Figures and legends

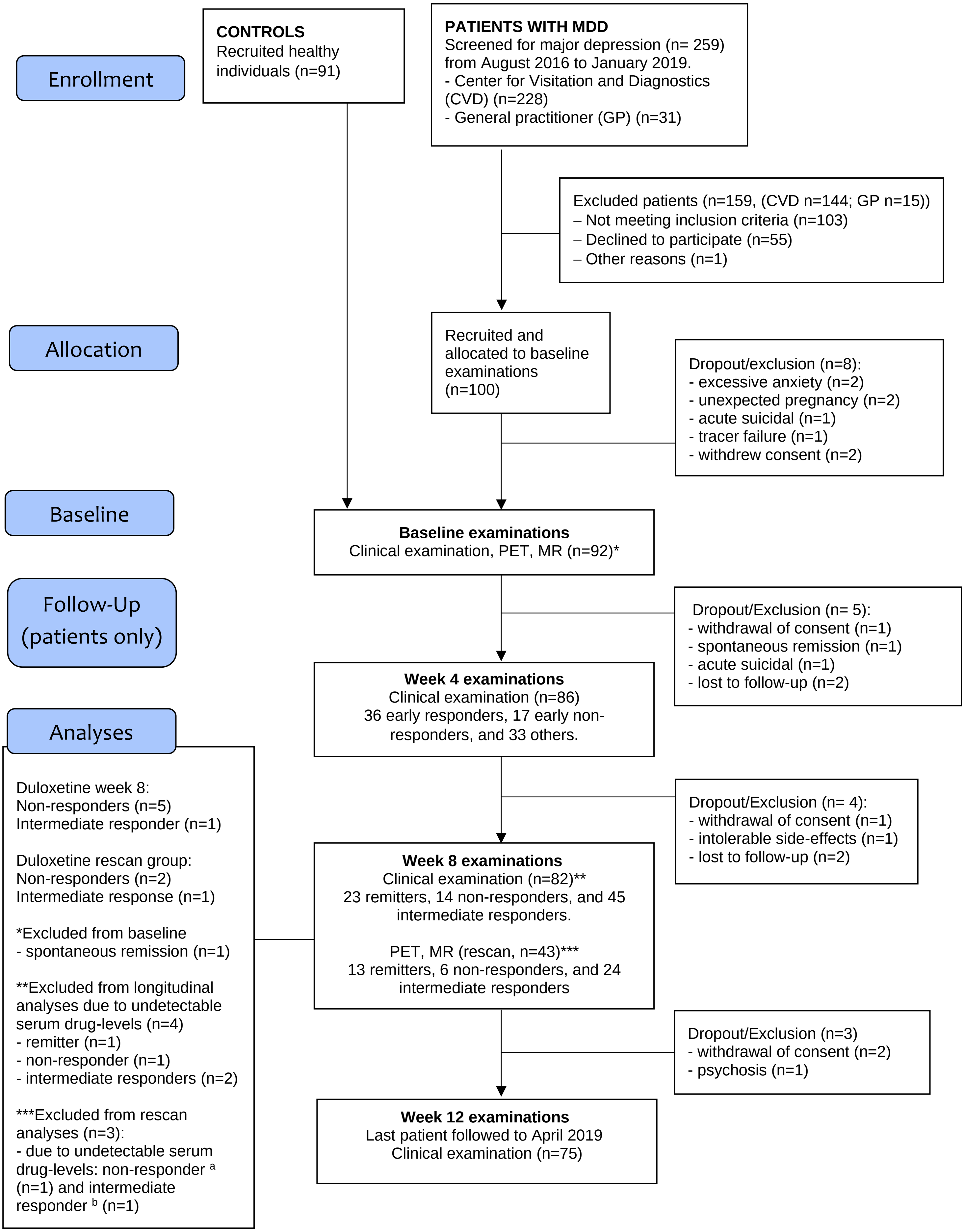

**Supplementary Figure 1. CONSORT flow diagram over study participants**. *One patient was excluded from baseline analyses because of spontaneous remission. **Four patients were excluded from the analyses at week 4, 8, and 12 because of undetectable serum-levels of escitalopram or duloxetine. *** Three patients were excluded from rescan analyses because of scanner failure and undetectable serum-levels of [a] duloxetine or [b] escitalopram.

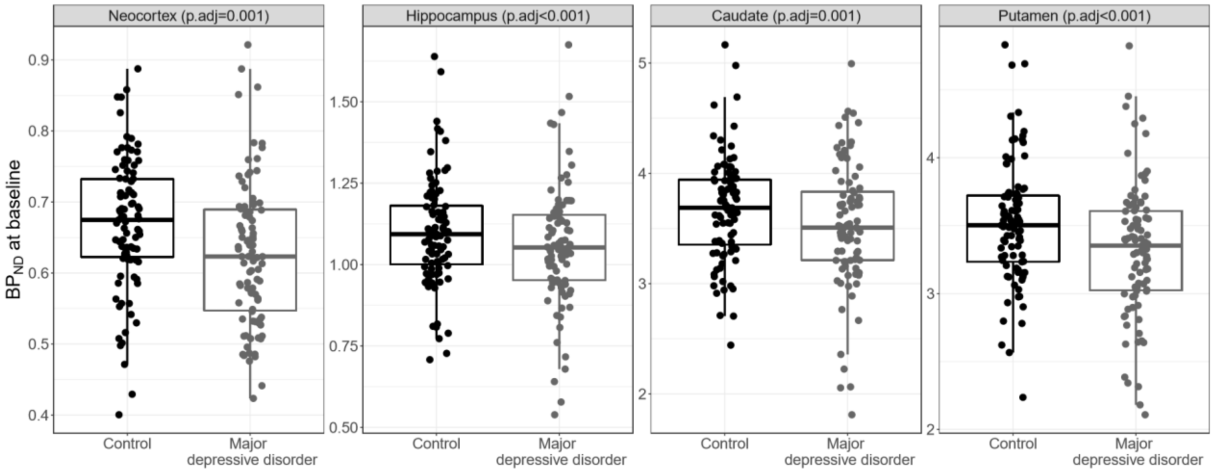


**Supplementary Figure 2. Scatter plot of regional cerebral baseline 5-HT$_4$R binding in patients with MDD and controls**.

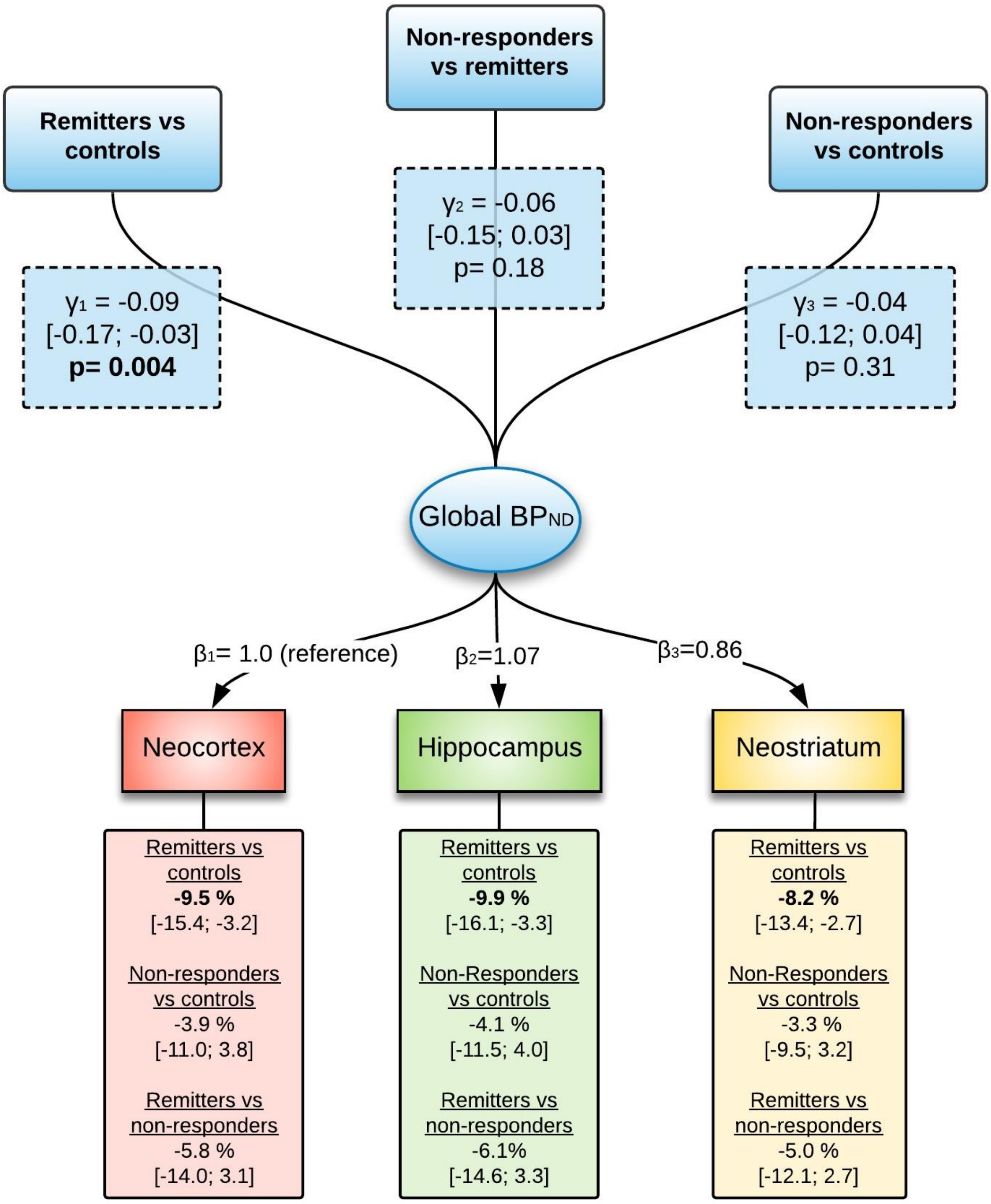


**Supplementary Figure 3. Latent variable model of differences in 5-HT$_4$R binding at baseline in controls and MDD, according to treatment response at week 8.** $\gamma$ is the effect of response-status on the global (log-transformed) $BP_{ND}$, $\beta$ is the regional loading. The lower boxes indicate, for each brain region, percentage differences in baseline 5-HT$_4$R binding between each response group and controls (p-values and confidence intervals are adjusted for 3 comparisons). The model includes the following covariates: age, sex, 5-HTTLPR gene-status, MR-scanner type and injected mass/kg.

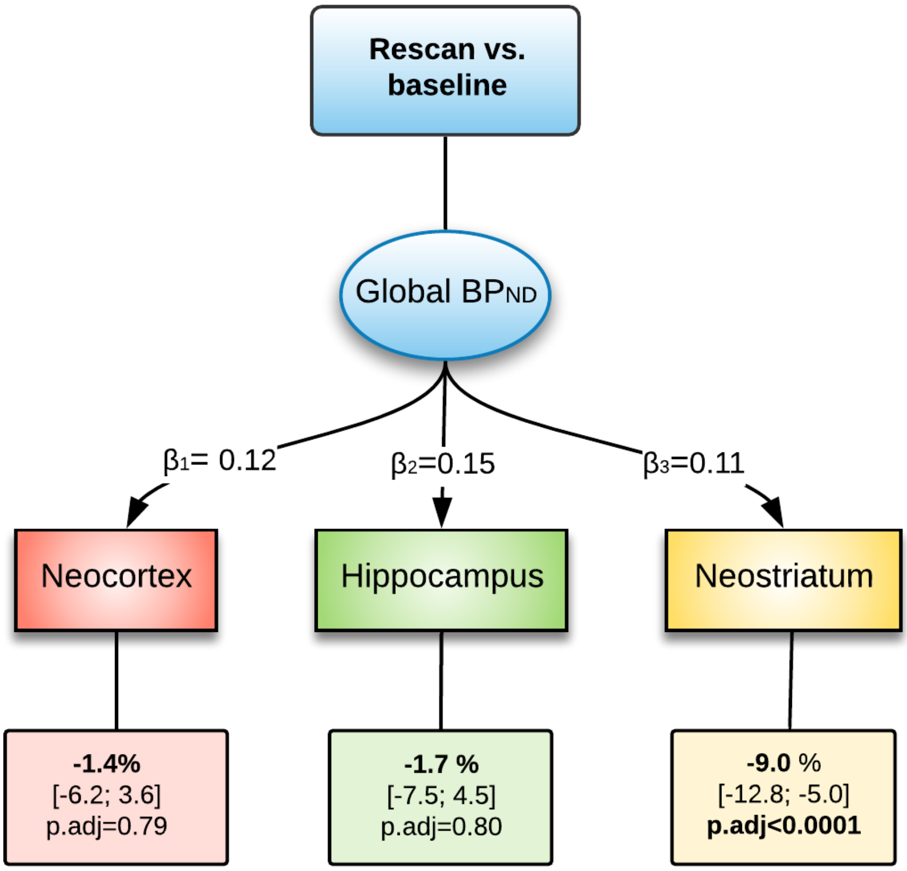


**Supplementary Figure 4. Latent variable model of changes in 5-HT$_4$R BP$_{ND}$ from baseline to rescan at week 8.** β is the regional loading. The lower boxes indicate, for each brain region, the percentage difference in 5-HT$_4$R binding from baseline to rescan. p-values and confidence intervals are adjusted for multiple comparisons. Injected SB207145 (mass/kg) was included in the model as a covariate.

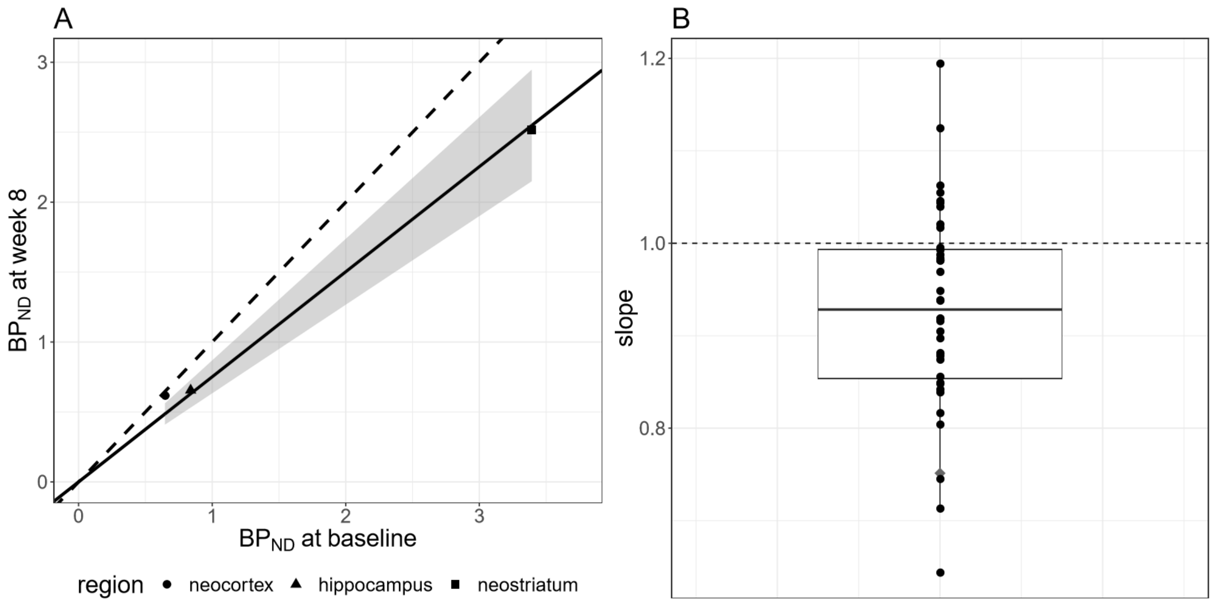


**Supplementary Figure 5. Left panel:** estimated regression line (black line) between the $BP_{ND}$ at baseline and at week 8 across three brain regions for a representative patient. The shaded area indicates the 95% confidence interval for the regression line and the dotted line the identity line (values below the dashed line indicates a decrease in $BP_{ND}$). **Right panel:** Boxplot with dots representing the individually estimated slopes of the regression line.

**Supplementary text for Supplementary Figure 5.**

In a post hoc analysis, we were able to replicate the findings from Haahr et al [17]. Using the same analysis strategy, we found a decrease in global binding (-0.07, 95% CI [-0.11 to -0.04], p=<0.001), see Supplementary Figure 5. Since we adapted a naturalistic design, we cannot directly know to what extent the decline in neostriatum 5-$HT_4$R binding is any larger than that seen in healthy controls [17], but the observation is a replication and extension of these prior findings and supports that long-term SSRI intervention increases brain serotonin levels also in patients with MDD.
